# Stroke Systems of Care: Evaluating Disparities after Stroke Legislation Implementation in Rhode Island

**DOI:** 10.64898/2025.12.12.25342179

**Authors:** Abigail A. Teshome, Joshua Feler, Mazen Taman, Carl Porto, Elizabeth Perelstein, Mark Cort, Elias Shaaya, Krisztina Moldovan, Radmehr Torabi, Dylan N. Wolman, Mahesh V. Jayaraman, Jeffrey Proulx

## Abstract

**Introduction:** The Rhode Island Stroke Prevention and Treatment Act aimed to establish a standardized system of care for stroke patients through designated stroke centers, revise emergency medical services (EMS) protocols, and establish a centralized stroke database. While Stroke Systems of Care (SSOC) laws have proven effective in reducing overall stroke mortality, their benefits among racial minorities and economically disadvantaged groups remain unclear. This study investigates the impact of the Act on stroke mortality rates across different racial groups.

**Methods:** Data from the Rhode Island Department of Labor and Training, CDC Atlas of Heart Disease and Stroke, and U.S. Census Bureau for 2005–2019 were analyzed. A difference-in-difference analysis compared mortality rates among White, Black, and Hispanic residents before and after the Act’s implementation. Trends were further validated using a mixed-effects model.

**Results:** Following SSOC implementation, the White group had significant reductions in ischemic stroke mortality (-5.38 per 100,000, 95% CI [-6.63, -4.12]; P<0.001). Analysis did not identify a difference in ischemic stroke mortality in the Hispanic (-2.67, 95% CI [-1.49, 6.83]; P=0.174) or Black group (4.69, 95% CI [-25.40, 16.01]; P=0.638). After Act-driven field triage implementation, there was a decline in all-stroke mortality among Black (-15.9 per 100,000, 95% CI [-30.52, -1.19]; P=0.035) and Hispanic (-7.07 per 100,000, 95% CI [-11.81, -2.32]; P=0.005) groups. However, the Black group maintained higher all-stroke mortality compared to the White population (RR 1.34, 95% CI [1.27, 1.41]; P<0.001).

**Conclusion:** While SSOC significantly reduced overall stroke mortality in Rhode Island, the benefits were not equitably distributed across racial groups. These findings underscore the need for future stroke legislation to incorporate culturally competent, evidence-based strategies addressing social determinants of health to reduce disparities and ensure equitable care.

## Introduction

Stroke Systems of Care (SSOC) laws are intended to optimize stroke treatment by coordinating the four major components of stroke care as defined by the Centers for Disease Control: prehospital patient assessment, emergency transport, acute treatment, and rehabilitation.^1^ Currently, 41 states in the U.S. have stroke legislation, though their contents are heterogenous; nearly half lack efficient emergency medical services (EMS) protocols, delaying access to care and increasing the risk of major disability and mortality.^2,3^ States with stroke legislation demonstrate lower age-adjusted stroke mortality, a greater number of certified stroke centers, better access to primary stroke centers (PSCs), and lower hospital costs.^3,4^ Despite these findings, there is limited research on the effectiveness of stroke policies among racial minorities. Communities of color, who often have greater prevalence of stroke risk factors, also experience barriers to acute stroke care, including lower EMS utilization, treatment delays, and lower likelihood of transfer to endovascular-capable or other centers.^5,6^

The Rhode Island Stroke Prevention and Treatment Act of 2009 sought to establish a standardized and comprehensive system of care for stroke patients in the state of Rhode Island. Key components of the Act included:

1. Designation of seven PSCs via Joint Commission (JC) certification
2. Designation of one acute ready stroke center (ARSC) adhering to American Heart Association/American Stroke Association (AHA/ASA) guidelines
3. Development of a statewide stroke database
4. Review and revision of EMS stroke protocols as a standardized assessment tool for all licensed EMS providers
5. Initiation of an updated EMS data collection system consistent with nationally recognized stroke treatment guidelines^7^

This Act was amended in 2015 following evidence supporting mechanical thrombectomy as standard of care for large vessel occlusion (LVO). This amendment called for:

1. Annually-reviewed pre-hospital care protocols including plans for triage to the closest CSC or PSC as appropriate (patients with suspected LVO based on field severity assessment to be triaged directly to a CSC)
2. Collaboration with the JC, AHA, and ASA for the development of a certification program for CSCs and PSCs
3. Participation with nationally recognized data set platforms and transmission of data to the statewide stroke registry
4. Establishment of reporting requirements to ensure uniformity and integrity of data submitted to the statewide registry^8^
5. A standardized inter-facility transfer mechanism between PSCs and the state’s only CSC
6. Modified regulations to allow easier transport of patients receiving intravenous thrombolytics, to facilitate interfacility transfer (“drip and ship”)

This study aims evaluates how SSOC law implementation has impacted stroke mortality rates among racial and ethnic minority populations in Rhode Island.

## Methods

Publicly available data from the Rhode Island Department of Labor and Training^9^ were collected to capture total population counts stratified by race for each county in Rhode Island between 2005 and 2019, reflecting four years before the implementation of the SSOC until four years after passage of the amendment and field triage implementation. Epidemiological data from the CDC Atlas of Heart Disease and Stroke^10^ were analyzed to assess stroke prevalence and mortality rates for ischemic stroke, hemorrhagic stroke, and all stroke, measured per 100,000 individuals aged 35 and older (35+) across White, Black, and Hispanic populations. Additionally, population age and gender estimates were collected from the US Census Bureau^11^ to account for population changes over time.

A difference-in-difference analysis was conducted to compare stroke mortality rates among White, Black, and Hispanic residents before and after the Act’s implementation in 2009, as well as following the passage of amendments in 2015. The pre-policy group included inclusively defined ranges from 2005-2007, 2006-2008, 2007-2009, and 2008-2010, while the post-policy group included ranges from 2009-2011, 2010-2012, 2011-2013, and 2012-2014. The post-triage group was 2014-2016, 2015-2017, 2016-2018, and 2017-2019. Analyses of the post-triage cohort were conducted in two contexts: (1) relative to the full pre-triage population which encompassed both pre- and post-policy periods, and (2) relative to the post-policy period alone. A mixed effects model assessed the relationship between county-level racial demographics and mortality rates from ischemic, hemorrhagic, and all strokes. Fixed effects included binary values for policy implementation or amendment passage and their interaction with race, while county was treated as a random effect to account for variation in baseline mortality between counties. Statistical analysis was performed in RStudio version 4.4.1.

## Results

### Epidemiological Landscape of Rhode Island

At the beginning of the study period, Rhode Island had 1,060,196 residents, with over half residing in Providence County (630,186). When isolating 35+ patients, there were 574,626 residents in the state with 341,560 in Providence County. **(Figure 1A)** This county houses the state’s only CSC as well as four of the seven designated PSCs and had the highest proportion of Black and Hispanic residents (9.2% and 16.5% respectively). In the same period, the least racially diverse county that year was Bristol County, which had a 35+ population of 27,175 (1.1% Black, 1.6% Hispanic) and no CSC designation. Over the study period, the total population in Rhode Island increased by 3.2%, and the 35+ population grew by 3.9%. Providence County had a 35+ population of 341,851, which was 10.3% Black and 17.5% Hispanic. Bristol County remained the least racially diverse county with 1.2% Black and 2.3% Hispanic representation. **(Figure 1B)**

**Figure 1.**
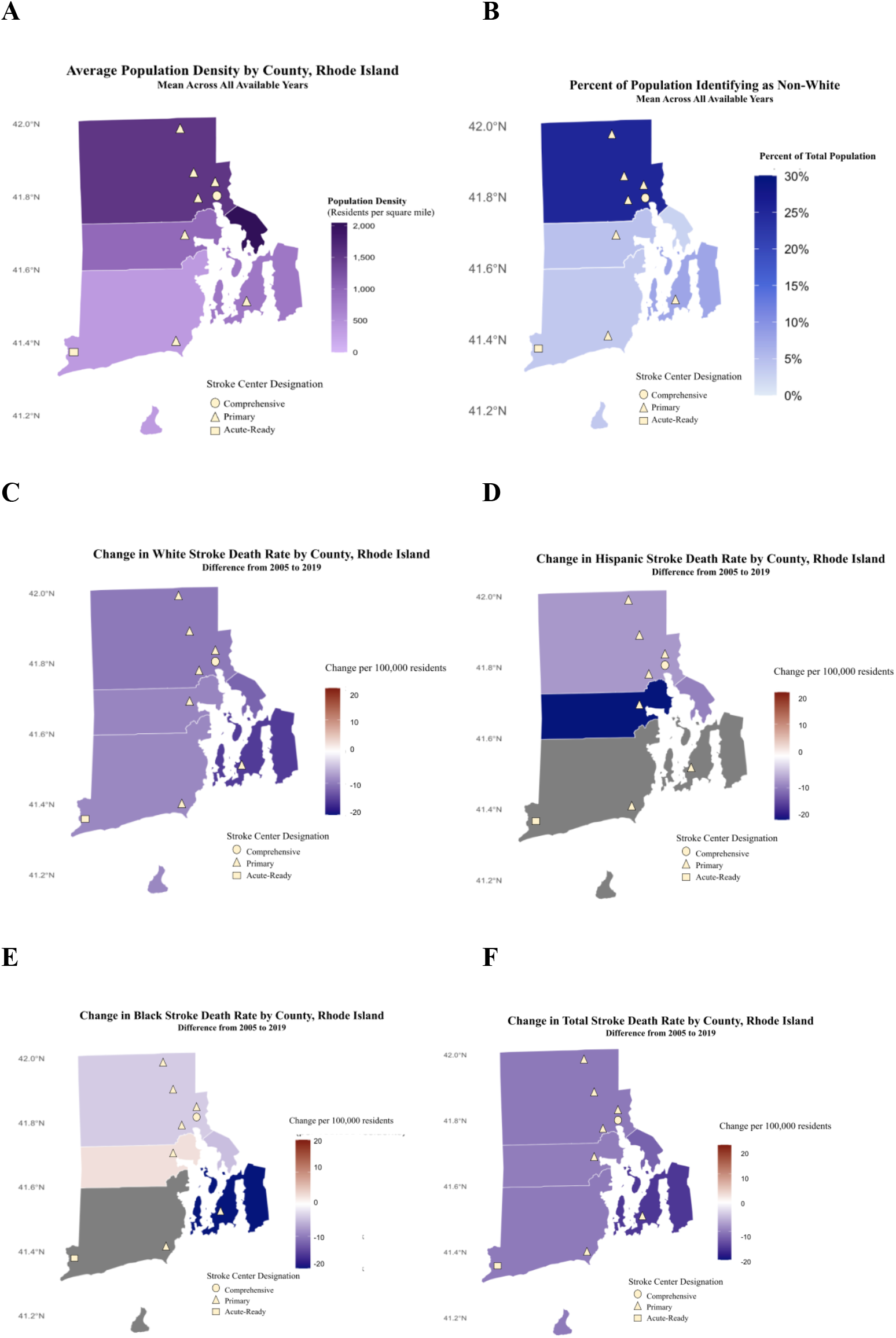
Heat map depicting **(A)** the average population density from 2005 to 2019 by Rhode Island County Providence County demonstrated the highest total population, but Bristol County had the highest population density across all years. Bristol was the only county without a designated stroke center after Act implementation. **(B)** The proportion of Non-White residents by county. Providence county had the highest proportion of Non-White residents and had the most amount of designated stroke centers, including the designation of the only comprehensive center. Change in **(C)** White and **(D)** Hispanic stroke death over the study period was negative in all counties. Change in **(E)** Black stroke death over the study period was positive in Kent County, and otherwise negative. **(F)** Change in the total population had similar trends to the White subgroup. Overlaid shapes represent the designation of stroke centers with the passage of the Act or amendment. Gray indicates insufficient data.

### Implementation of the Stroke Prevention and Treatment Act of 2009

After SSOC implementation, all-stroke mortality decreased statewide by 8.56 deaths per 100,000 (95% CI [-10.56, -6.56]; P<0.001). Likewise, White and Hispanic subgroups demonstrated a decline; however, the Black population was not identified to have changed. **(Table 1A)** When stratifying by stroke subtype, only the White population experienced declines in ischemic stroke mortality (-5.38 per 100,000, -6.63, -4.13]; P<0.001). Mortality among Black and Hispanic residents remained unchanged across both subtypes. **(Table 1A)**

**Table 1.**
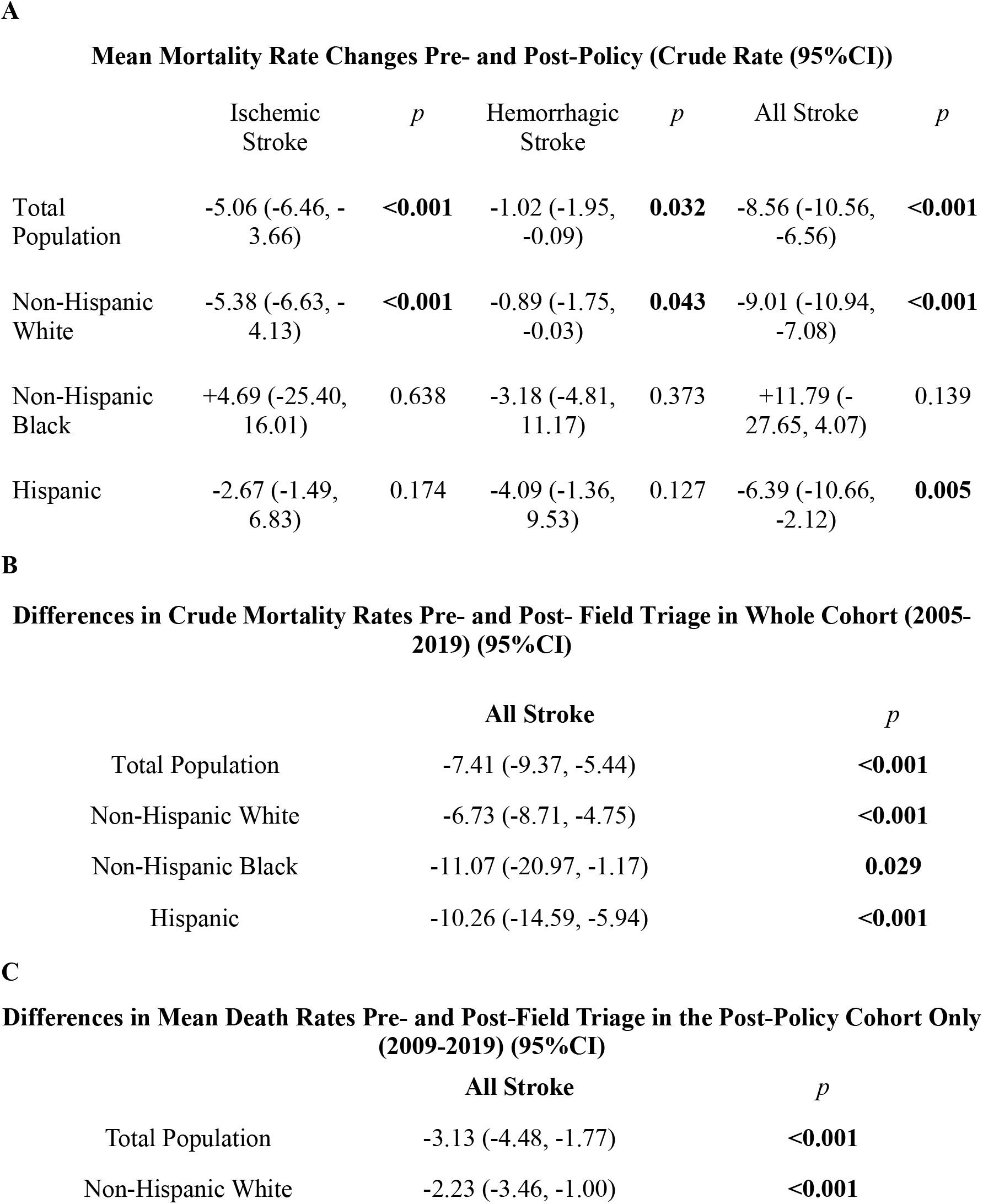
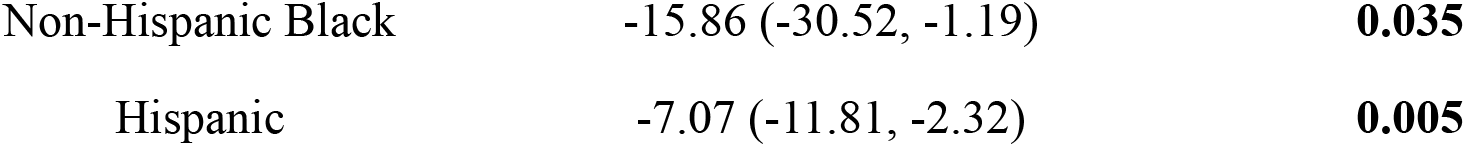
Difference-in-difference analysis of stroke mortality rates among the total population and stratified by race. (**A)** After policy implementation, all stroke mortality only decreased in the total, White, and Hispanic population. The Black population demonstrated an increase in all stroke mortality during this period. **(B)** In the four years before and after field triage implementation in 2015, the Black and Hispanic populations demonstrated decreases in all stroke mortality while there was no change in the White population. **(C)** When analyzing the entire sample and categorizing pre- and post-field triage eras, all groups demonstrated a decline in all stroke mortality.

**Mean Mortality Rate Changes Pre- and Post-Policy (Crude Rate (95%CI))**
|  | Ischemic<br>Stroke | *p* | Hemorrhagic<br>Stroke | *p* | All Stroke | *p* |
| --- | --- | --- | --- | --- | --- | --- |
| Total<br>Population | -5.06 (-6.46, -<br>3.66) | **<0.001** | -1.02 (-1.95,<br>-0.09) | **0.032** | -8.56 (-10.56,<br>-6.56) | **<0.001** |
| Non-Hispanic<br>White | -5.38 (-6.63, -<br>4.13) | **<0.001** | -0.89 (-1.75,<br>-0.03) | **0.043** | -9.01 (-10.94,<br>-7.08) | **<0.001** |
| Non-Hispanic<br>Black | +4.69 (-25.40,<br>16.01) | 0.638 | -3.18 (-4.81,<br>11.17) | 0.373 | +11.79 (-<br>27.65, 4.07) | 0.139 |
| Hispanic | -2.67 (-1.49,<br>6.83) | 0.174 | -4.09 (-1.36,<br>9.53) | 0.127 | -6.39 (-10.66,<br>-2.12) | **0.005** |

**B** **Differences in Crude Mortality Rates Pre- and Post- Field Triage in Whole Cohort (2005-2019) (95%CI)**
|  | All Stroke | *p* |
| --- | --- | --- |
| Total Population | -7.41 (-9.37, -5.44) | **<0.001** |
| Non-Hispanic White | -6.73 (-8.71, -4.75) | **<0.001** |
| Non-Hispanic Black | -11.07 (-20.97, -1.17) | **0.029** |
| Hispanic | -10.26 (-14.59, -5.94) | **<0.001** |

**Table 1.** Difference-in-difference analysis of stroke mortality rates among the total population and stratified by race.
|  | All Stroke | *p* |
| --- | --- | --- |
| Total Population | -3.13 (-4.48, -1.77) | **<0.001** |
| Non-Hispanic White | -2.23 (-3.46, -1.00) | **<0.001** |
| Non-Hispanic Black | -15.86 (-30.52, -1.19) | **0.035** |
| Hispanic | -7.07 (-11.81, -2.32) | **0.005** |

A mixed effects model revealed that the pre-policy mortality rate from all strokes among Black residents was higher than White residents (RR 1.17, 95% CI [1.07, 1.27]; *P*<0.001), while Hispanic residents had lower pre-policy mortality from all strokes (RR 0.83, 95% CI [0.76, 0.92]; *P*<0.001) compared to White residents. Compared to the White population, the Black population demonstrated a relative increase in all-stroke mortality during this period (RR 1.30, 95% CI [1.16, 1.45]; *P*<0.001). **(Table 2A)**

**Table 2.**
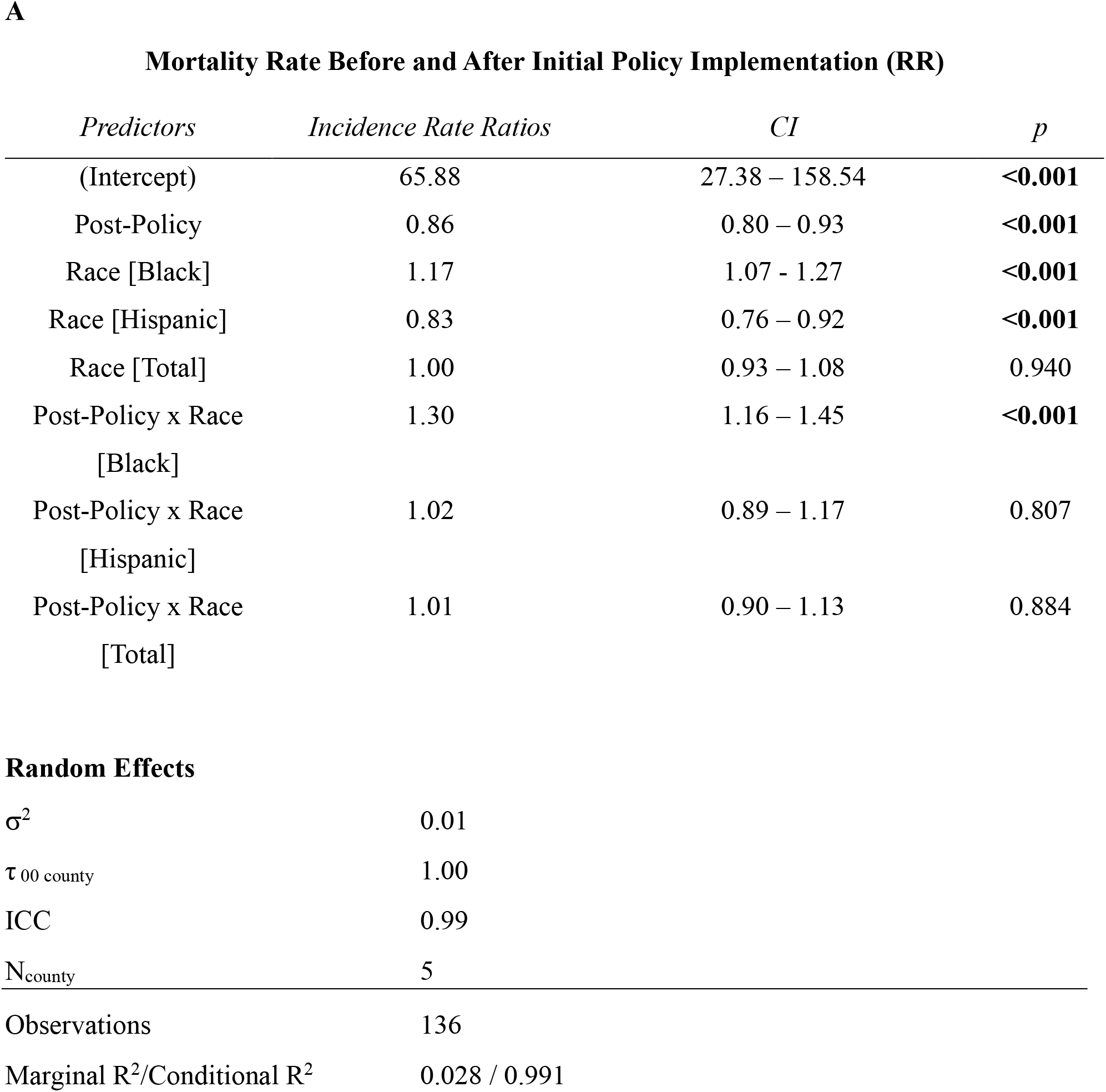
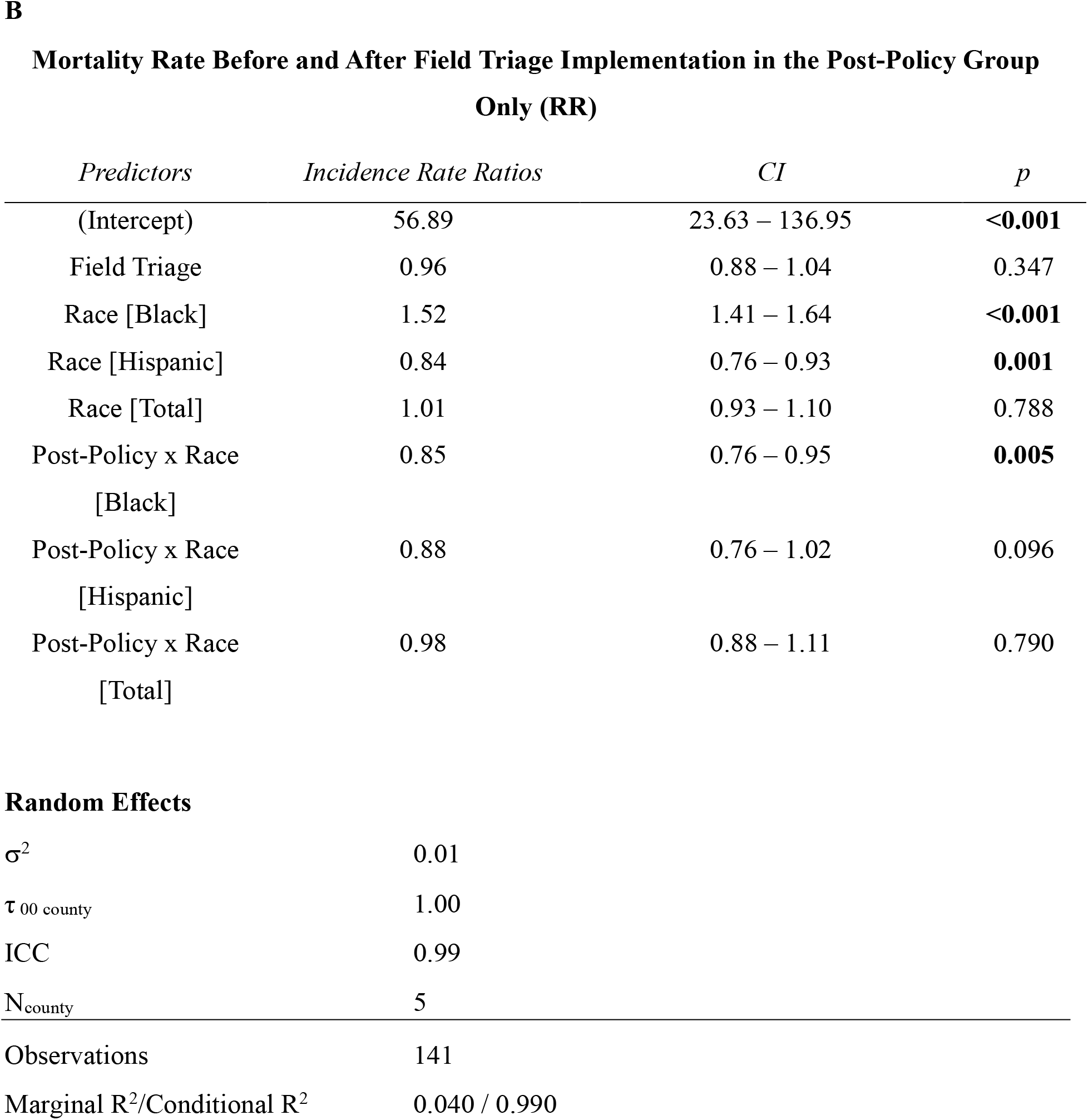
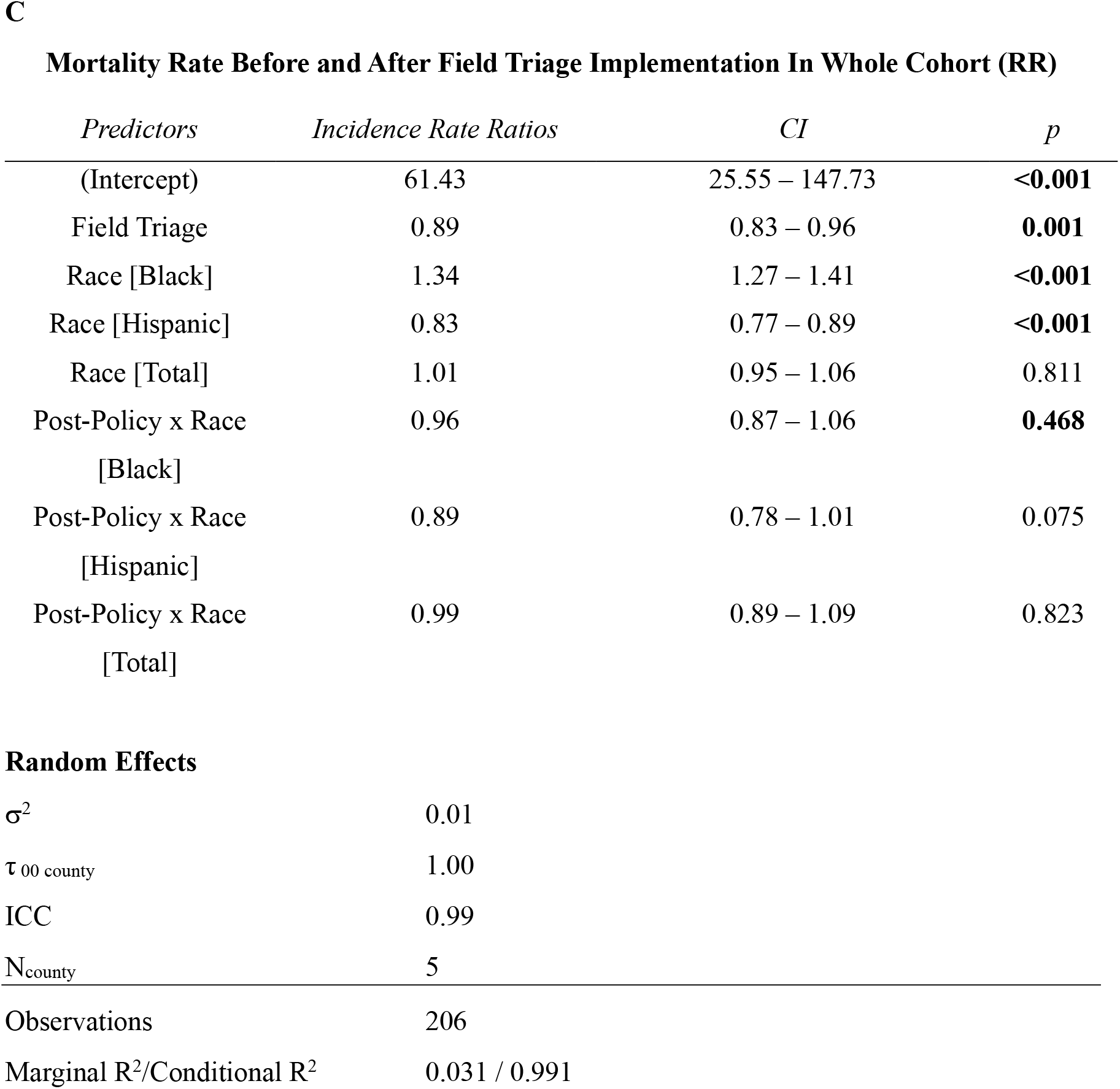
Results from amixed effects model in which the intercept is defined as the reference (White) group. **(A)** When analyzing the four years before (2005-2009) and after (2009-2014) policy implementation, the Black subgroup demonstrated a higher pre-policy mortality rate than the White population. This trend followed into the post-policy period. The Hispanic group had a lower pre-policy mortality rate than the White group, but analysis showed no difference in the change in mortality after policy implementation between the two groups. **(B)** When comparing the sample before (2011-2015) and after (2015-2019) thrombectomy implementation, the Black population demonstrated a higher mortality compared to the White group while the Hispanic population had a lower mortality rate during the pre-thrombectomy period. After thrombectomy implementation, the Black group demonstrated a relative decrease in mortality. **(C)** When analyzing the entire pre-thrombectomy group (2005-2015) and comparing it against the post-thrombectomy group (2015-2019), the Hispanic population had a lower pre-thrombectomy mortality rate while the Black population had a higher mortality rate during this period relative to the White group. No differences were found in the mortality rate change before and after thrombectomy implementation.

| *Predictors* | *Incidence Rate Ratios* | *CI* | *p* |
| --- | --- | --- | --- |
| (Intercept) | 65.88 | 27.38 – 158.54 | **<0.001** |
| Post-Policy | 0.86 | 0.80 – 0.93 | **<0.001** |
| Race [Black] | 1.17 | 1.07 - 1.27 | **<0.001** |
| Race [Hispanic] | 0.83 | 0.76 – 0.92 | **<0.001** |
| Race [Total] | 1.00 | 0.93 – 1.08 | 0.940 |
| Post-Policy x Race<br>[Black] | 1.30 | 1.16 – 1.45 | **<0.001** |
| Post-Policy x Race<br>[Hispanic] | 1.02 | 0.89 – 1.17 | 0.807 |
| Post-Policy x Race<br>[Total] | 1.01 | 0.90 – 1.13 | 0.884 |

**Random Effects**
|  |  |
| --- | --- |
| $\sigma^2$ | 0.01 |
| $\tau_{00 \text{ county}}$ | 1.00 |
| ICC | 0.99 |
| $N_{\text{county}}$ | 5 |
| Observations | 136 |
| Marginal R<sup>2</sup>/Conditional R<sup>2</sup> | 0.028 / 0.991 |

**Mortality Rate Before and After Field Triage Implementation in the Post-Policy Group Only (RR)**
| *Predictors* | *Incidence Rate Ratios* | *CI* | *p* |
| --- | --- | --- | --- |
| (Intercept) | 56.89 | 23.63 – 136.95 | **<0.001** |
| Field Triage | 0.96 | 0.88 – 1.04 | 0.347 |
| Race [Black] | 1.52 | 1.41 – 1.64 | **<0.001** |
| Race [Hispanic] | 0.84 | 0.76 – 0.93 | **0.001** |
| Race [Total] | 1.01 | 0.93 – 1.10 | 0.788 |
| Post-Policy x Race<br>[Black] | 0.85 | 0.76 – 0.95 | **0.005** |
| Post-Policy x Race<br>[Hispanic] | 0.88 | 0.76 – 1.02 | 0.096 |
| Post-Policy x Race<br>[Total] | 0.98 | 0.88 – 1.11 | 0.790 |

**Random Effects**
|  |  |
| --- | --- |
| $\sigma^2$ | 0.01 |
| $\tau_{00 \text{ county}}$ | 1.00 |
| ICC | 0.99 |
| $N_{\text{county}}$ | 5 |
| Observations | 141 |
| Marginal R<sup>2</sup>/Conditional R<sup>2</sup> | 0.040 / 0.990 |

**Mortality Rate Before and After Field Triage Implementation In Whole Cohort (RR)**
| *Predictors* | *Incidence Rate Ratios* | *CI* | *p* |
| --- | --- | --- | --- |
| (Intercept) | 61.43 | 25.55 – 147.73 | **<0.001** |
| Field Triage | 0.89 | 0.83 – 0.96 | **0.001** |
| Race [Black] | 1.34 | 1.27 – 1.41 | **<0.001** |
| Race [Hispanic] | 0.83 | 0.77 – 0.89 | **<0.001** |
| Race [Total] | 1.01 | 0.95 – 1.06 | 0.811 |
| Post-Policy x Race<br>[Black] | 0.96 | 0.87 – 1.06 | **0.468** |
| Post-Policy x Race<br>[Hispanic] | 0.89 | 0.78 – 1.01 | 0.075 |
| Post-Policy x Race<br>[Total] | 0.99 | 0.89 – 1.09 | 0.823 |

**Table 2.** Results from amixed effects model in which the intercept is defined as the reference (White) group.
|  |  |
| --- | --- |
| $\sigma^2$ | 0.01 |
| $\tau_{00 \text{ county}}$ | 1.00 |
| ICC | 0.99 |
| $N_{\text{county}}$ | 5 |
| Observations | 206 |
| Marginal R<sup>2</sup>/Conditional R<sup>2</sup> | 0.031 / 0.991 |

### Implementation of Field Triage and Amendment of the Act

Across the entire pre-vs post-field triage period, all groups demonstrated significant reductions in all stroke mortality. **(Figure 1)** A mixed effects model revealed that the Black and Hispanic populations showed an insignificant change in mortality when compared to the White subgroup. **(Table 2B)** When analyzing those in the post-policy cohort only, all groups demonstrated declines in mortality **(Table 2C)**. Despite this decrease, Black residents continued to have significantly higher all stroke mortality compared with White residents (RR 1.52, 95% CI [1.41, 1.64]; P<0.001).

## Discussion

This study evaluated how the Rhode Island Stroke Prevention and Treatment Act of 2009, its amendment, and the rise of mechanical thrombectomy affected the landscape of stroke care and racial disparities in stroke mortality. While the implementation of the initial Act in 2009 was associated with lower overall stroke mortality statewide, these benefits were not equally distributed across racial groups. White residents experienced significant mortality reductions across stroke types, whereas Black and Hispanic residents demonstrated limited or no improvement. These findings align with broader trends of improved stroke outcomes following SSOC implementation but also reveal persistent racial disparities in outcomes which have not been highlighted in literature to date. Implementation of field triage for suspected LVO was associated with decreased mortality rates among the Black and Hispanic populations. Despite this, the mortality rate among the Black population remained higher than any other race across time periods. **(Figure 1E)**

Policy interventions associated with improved stroke outcomes include creation of an SSOC task force, a statewide continuous quality improvement database, tiered stroke center designation, EMS stroke assessment tools, and standardized transport protocols.^1^ However, these largely address acute care processes, while the persistent disparities observed here may reflect chronic social and structural factors. There is evidence that the tiered stroke center designation emphasis in the 2015 amendment may be associated with the decrease in mortality among Black and Hispanic groups. Direct transfer to thrombectomy-capable centers when LVO is suspected has been linked to shorter length of hospital stay and reduced mortality in both urban and rural settings.^12-14^

Despite the decline in mortality among the Black group, it remained the highest in all-stroke mortality. Current evidence suggests that while certification of various stroke center types is associated with increased admission rates to certified centers, there is no change in thrombolysis or thrombectomy rates for Black patients living near newly certified centers.^15^ In addition, primary language, socioeconomic status, and health literacy strongly influence time to presentation, eligibility for reperfusion therapies, and functional recovery.^16-18^ Non-English-speaking patients may present with greater stroke severity and worse outcomes, likely due to barriers to effective communication in acute emergencies and reduced preventive care access.^16,19^ According to Migration Policy Institute, more than 20,000 Rhode Island residents speak African and Caribbean languages, of which upwards of 36% identify as having a non-proficient understanding of English. Additionally, among the estimated 116,674 Spanish speaking adults in the state, 50.4% are proficient in English.^20^

Despite the increased proportion of racial minority representation in urban tracts with higher stroke center density, time to treatment may be increased among racial minorities.^6,21,22^ Lower utilization of EMS, less accurate prehospital recognition of stroke by EMS, and differences in the perception of stroke severity on evaluation by medical providers may contribute to this disparity.^5,21,23,24^ These findings underscore that improvements in system infrastructure alone are insufficient. Addressing the root causes of disparity will require culturally competent outreach and equity-focused system design. In Rhode Island, programs such as Progreso Latino and Dorcas International enhance access to language education, employment opportunities, and healthcare navigation, factors linked to improved health and stroke outcomes.^25,26^ Additionally, integrating community health workers and embedding culturally relevant education and support into care protocols may effectively bridge gaps, ensuring that health care is inclusive, accessible, and tailored to the specific needs of all populations.^27,28^

Although increasing the number of designated stroke centers improves access, many racially diverse communities already live near such centers, suggesting that proximity may not equate to accessibility. Our results emphasize that stroke legislation may not address key social determinants of health. Policymakers may consider prioritizing diversity in healthcare leadership, equity-focused EMS training, and funding for outreach initiatives that build trust and reduce barriers to care. Embedding equity metrics within SSOC evaluation frameworks may help ensure that legislative successes translate into equitable health outcomes across racial and ethnic groups.

### Limitations

A key limitation of this study is the absence of data for Asian and Native American populations, as the CDC Atlas of Heart Disease and Stroke does not report mortality rates for these groups in Rhode Island due to small population sizes. This omission underscores the need for more comprehensive data collection to understand how legislation impacts all racial and ethnic communities. In addition, county-level analysis may obscure within-county variation, warranting future analyses at the ZIP code or census tract level. Expanding metrics to include quality-of-life and functional recovery outcomes, as well as exploring interventional strategies such as mobile stroke units and telemedicine, could better inform efforts to create an equitable and effective stroke system of care. Future research should evaluate stroke interventions such as thrombolysis and thrombectomy while examining biological, behavioral, and social determinants of outcomes. The extent to which our findings are due to the designation of and updated triage protocols to CSCs independent of thrombectomy use remains unclear.

## Conclusions

While the SSOC significantly advanced stroke care in Rhode Island, its impact on reducing mortality was disproportionate, with White residents benefiting most and Black and Hispanic populations seeing limited gains. The introduction of CSC designation and field triage, pre-hospital stroke protocols, and mechanical thrombectomy were associated with reductions in mortality across racial groups. However, persistently high mortality among Black patients underscores the need for targeted interventions. Addressing structural inequities will require a multifaceted, culturally competent, evidence-based approach tailored to the needs of diverse communities. This study provides a critical foundation for future research and policy development to promote equity in stroke care and outcomes.

## Data Availability

Data is from publicly available datasets including US Census, RI Department of Labor and Training, and CDC Atlas of Heart Disease and Stroke.

https://dlt.ri.gov

https://www.cdc.gov/heart-disease-stroke-atlas/about/index.html

https://www.census.gov

